# Exploratory data on the clinical efficacy of monoclonal antibodies against SARS-CoV-2 Omicron Variant of Concern

**DOI:** 10.1101/2022.05.06.22274613

**Authors:** Fulvia Mazzaferri, Massimo Mirandola, Alessia Savoldi, Pasquale De Nardo, Matteo Morra, Maela Tebon, Maddalena Armellini, Giulia De Luca, Lucrezia Calandrino, Lolita Sasset, Denise D’Elia, Emanuela Sozio, Elisa Danese, Davide Gibellini, Isabella Monne, Giovanna Scroccaro, Nicola Magrini, Annamaria Cattelan, Carlo Tascini, MANTICO Working Group, Evelina Tacconelli

## Abstract

**Background:** Recent in-vitro data have shown that the activity of monoclonal antibodies (mAbs) targeting SARS-CoV-2 varies according to the Variant of Concern (VOC). No studies have compared the clinical efficacy of different mAbs against Omicron VOC.

**Methods:** The MANTICO trial is a non-inferiority randomised controlled trial comparing the clinical efficacy of bamlanivimab/etesevimab, casirivimab/imdevimab, and sotrovimab in outpatients aged 50 or older with early COVID-19. As the patient enrolment was interrupted for possible futility after the onset of the Omicron wave, the analysis was performed according to the SARS-CoV-2 VOC. The primary outcome was COVID-19 progression (hospitalisation, need of supplemental oxygen therapy, or death through day 14). Secondary outcomes included the time to symptom resolution, assessed using the product-limit method. Kaplan-Meier estimator and Cox proportional hazard model were used to assess the association with predictors. Log rank test was used to compare survival functions.

**Results:** Overall, 319 patients were included. Among 141 patients infected with Delta, no disease progression was recorded and the time to symptom resolution did not differ significantly between treatment groups (Log-rank Chi-square 0.22, *p* 0.895). Among 170 patients infected with Omicron (80.6% BA.1, 19.4% BA.1.1), two disease progressions were recorded in the bamlanivimab/etesevimab group and the median time to symptom resolution was 5 days shorter in the sotrovimab group compared to bamlanivimab/etesevimab and casirivimab/imdevimab (HR 0.526 and HR 0.451, 95% CI 0.359 - 0.77 and 95% CI 0.303 - 0.669, *p* 0.001 and 0.0001, respectively).

**Conclusions:** These results confirm the in-vitro data of superiority of sotrovimab versus casirivimab/imdevimab and bamlanivimab/etesivamab in reducing the time to recovery in patients infected with Omicron BA.1 and BA.1.1, while no difference was detected in Delta infections. Casirivimab/imdevimab seems to maintain a role in preventing severe COVID-19 in the Omicron population. Adaptive clinical trials comparing mAbs by VOC should be pursued to promptly inform clinical recommendations.

**Funding:** This trial was funded by the Italian Medicines Agency (Agenzia Italiana del Farmaco, AIFA). The VOC identification was funded by the ORCHESTRA (Connecting European Cohorts to Increase Common and Effective Response to SARS-CoV-2 Pandemic) project, which has received funding from the European Union’s Horizon 2020 research and innovation programme under grant agreement No 101016167.

**Clinical trial number:** NCT05205759

## Introduction

Coronavirus disease 2019 (COVID-19), which is caused by severe acute respiratory syndrome coronavirus 2 (SARS-CoV-2), has spread globally and poses a major challenge to healthcare systems worldwide. A high incidence of hospitalisation and death due to COVID-19 has been reported among older patients and those with certain coexisting conditions, such as obesity, diabetes mellitus, cardiovascular disease, chronic obstructive pulmonary disease, and chronic kidney disease [1, 2]. The implementation of mass vaccination campaigns has markedly reduced the healthcare burden related to COVID-19. Nevertheless, SARS-CoV-2 vaccination rates differ considerably across countries and growing evidence suggests a reduced efficacy of vaccines against new viral Variants of Concern (VOC) [3-6].

Therapeutic agents directed against SARS-CoV-2 have been developed to prevent the disease progression, especially addressing high-risk groups of patients. Neutralizing monoclonal antibodies (mAbs) target the spike protein of SARS-CoV-2 that mediates viral entry into host cells [7]. Based on the results of randomised placebo-controlled trials showing the efficacy in preventing COVID-19 progression, drug regulatory authorities, such as the US Food and Drug Administration (FDA), the European Medicines Agency (EMA), and the Italian Medicines Agency (AIFA), had granted the emergency use authorization status for bamlanivimab 700 mg combined with etesevimab 1400 mg, casirivimab 600 mg combined with imdevimab 600 mg, and sotrovimab 500 mg to treat early COVID-19 in patients at high risk of progression [8-10]. To date, no randomised trials have compared the efficacy of these mAbs in preventing severe COVID-19.

This paper reports the results of the MANTICO trial, a non-inferiority randomised controlled trial comparing the clinical efficacy of routinely-used mAbs in a real-life setting of outpatients aged 50 or older with early mild-to-moderate COVID-19. The patient enrolment started in December 2021 and was interrupted after the publication of in-vitro evidence that two treatments under investigation (bamlanivimab/etesevimab and casirivimab/imdevimab) were not effective against the new emerging viral Omicron VOC [3-5]. The analysis is therefore restricted to 319 randomised patients, who were enrolled up to the interruption for possible futility, and was performed according to the SARS-CoV-2 VOC (Delta and Omicron).

## Methods

### Trial design

The trial was designed as a pragmatic, randomised, single-blind, non-inferiority, parallel group, multi-centre, controlled trial. Eligible subjects were outpatients aged 50 years or older, presenting with a positive test (either direct antigen or nucleic acid SARS-CoV-2) and mild to moderate COVID-19 symptoms within 4 days of the onset [11]. COVID-19 symptoms included cough, nasal congestion, sore throat, feeling hot or feverish, myalgia, fatigue, headache, anosmia/ageusia, nausea, vomiting, and/or diarrhoea [12]. Predefined exclusion criteria included a peripheral oxygen saturation level of 93% or less on room air, a respiratory rate of 30 or more breaths per minute, a heart rate of 125 or more beats per minute, and previous COVID-19 treatments with mAbs.

Participants were randomly assigned in a 1:1:1 ratio to receive a single intravenous infusion over a period of 1 hour, consisting of a combination of 700 mg of bamlanivimab and 1400 mg of etesevimab or 500 mg of sotrovimab or a combination of 600 mg of casirivimab and 600 mg of imdevimab. The study drugs were diluted to 250 mL with normal saline. Patients were masked to treatment group assignment. Randomisation was computer-generated in permuted blocks with a stratification based on site. The allocated drug was revealed to the investigator using an online randomisation module within the REDCap data management system [13].

The trial was conducted in accordance with the principles of the Declaration of Helsinki, the international ethical guidelines of the Council for International Organizations of Medical Sciences, the International Council for Harmonisation Good Clinical Practice guidelines, and applicable laws and regulations. All patients or their legally authorized representatives provided written informed consent. This study is registered with ClinicalTrials.gov, NCT05205759, where details on the sample size determination are provided.

### Outcomes

The composite primary outcome was the COVID-19 progression, defined as hospitalisation, need of supplemental oxygen therapy, or death from any cause through day 14. The presence of any of the three variables qualified the presence of the COVID-19 progression. Prespecified secondary outcomes were emergency department visits through day 28, all-cause mortality through day 28, duration of supplemental oxygen therapy, rate and duration of non-invasive ventilation and mechanical ventilation, and time to sustained patient-reported symptom resolution, which was defined as the absence of any symptom related to COVID-19 for at least 24 hours [14].

### Predictors

The main predictor was the treatment regimen randomised at enrolment (bamlanivimab/etesevimab, casirivimab/imdevimab, and sotrovimab). All patients were assessed at baseline for the following predictors to be tested for association with the time to symptom resolution: age, sex, body mass index, relevant comorbidities (diabetes for which medication was warranted, cardiovascular disease [hypertension, coronary artery disease, congestive heart failure], chronic kidney disease, chronic liver disease, chronic pulmonary disease, active cancer, transplant, and other immunocompromising conditions), SARS-CoV-2 serological status (anti-spike IgG), and SARS-CoV-2 vaccination status. The SARS-CoV-2 serological status was categorized as serum antibody-negative (if test results were negative), serum antibody-positive (if test results were positive), or other (inconclusive or unknown results). The SARS-CoV-2 vaccination status was categorised as not vaccinated, partial or complete primary COVID-19 vaccination series administered more than 180 days before the enrolment, complete primary COVID-19 vaccination series administered 180 days or less before the enrolment, and booster vaccination [6]. These categories were further collapsed as not vaccinated and partial or complete primary COVID-19 vaccination series administered more than 180 days before the enrolment versus complete primary COVID-19 vaccination series administered 180 days or less before the enrolment and booster vaccination.

### Procedures and tools

Outpatient visits were scheduled at baseline, 14±3 days and 30±3 days after the randomisation. Patients were considered lost to follow-up if they repeatedly did not participate in scheduled visits and could not be contacted by the investigators. Medical evaluation, vital signs recording, and laboratory tests were performed at each visit. If patients missed the visits, they were called by telephone to assess clinical conditions.

The SARS-CoV-2 serological status was assessed using LIAISON SARS-CoV-2 TrimericS IgG assay (DiaSorin), an indirect chemiluminescence immunoassay (CLIA) detecting IgG against the spike viral protein in its native trimeric conformation, which includes receptor-binding domain and N-terminal domain sites from the three subunit S1. According to the to manufacturer’s instructions, Binding Antibody Units (BAU)/mL ≥ 33.8 were considered positive for anti-trimeric spike protein specific IgG antibodies.

Nasopharyngeal swabs were processed using MagMAX Viral/Pathogen Nucleic Acid Isolation Kit and KingFisher automated extraction system (ThermoFisher Scientific). Viral RNA was detected using COVIDSeq amplicon-based Next Generation Sequencing Test combined with COVIDSeq V4 Primer Pool (Illumina, Inc.). Sequencing libraries were synthesized using automated Microlab STAR liquid handler (Hamilton Company). Pooled samples were quantified using Qubit 2.0 fluorometer (Invitrogen Inc.). Next generation sequencing was performed in 150 PE mode on NextSeq 550 Sequencing System (Illumina, Inc.) or MiSeq System (Illumina, Inc.) using the NextSeq 500/550 Mid Output Kit v2.5 or the Miseq Reagent Kit v3, respectively.

### Statistical analysis

For continuous variables, mean and standard deviation or median and IQR were calculated. For categorical variables, count and percentages were used. All outcome variables estimates were reported with 95% confidence interval (95% CI). Wilcoxon–Mann–Whitney test was used to compare independent groups. The association between categorical variables was assessed using the Fisher’s test. The product-limit method (Kaplan and Meier) was used to describe the time to symptom resolution. Kaplan-Meier estimator and Cox proportional hazard model were used to assess the bivariate association of independent variables with the time-dependent outcome. Kaplan-Meier curves were plotted to depict the association between each predictor and symptom persistence and the Log-rank test was used to compare survival functions. Predictors associated with the time to symptom resolution with a probability < 0.05 were considered significant. A two-sided test of less than 0.05 was considered statistically significant in all analyses. All statistical analyses were performed with the use of Stata Version 17.0 (College Station, TX: StataCorp LP).

## Results

The first patient was enrolled on December 9, 2021. Overall, 319 patients underwent randomisation by January 20, 2022, and were assigned to receive bamlanivimab/etesevimab (106 patients), sotrovimab (107 patients), or casirivimab/imdevimab (106 patients). No patients reported previous SARS-CoV-2 infections. No patients withdrew from the trial. VOC data were available for 311 patients: 170 (53.3%) were infected with Omicron and 141 (44.2%) with Delta. Eight (2.5%) patients were excluded from this analysis due to the lack of SARS-CoV-2 VOC identification. Baseline characteristics of the population by type of SARS-CoV-2 VOC are reported in Table 1.

**Table 1.** Baseline characteristics of the overall study population by type of Variant of Concern.

| Characteristic | Delta<br>N = 141 | Omicron<br>N = 170 | p value |
| --- | --- | --- | --- |
| **Sex (male) – n (%)** | 77 (54.61) | 101 (59.41) | 0.421 |
| **Age – median (interquartile range) [range]** | 65.7 (15.4) [50-92] | 64.5 (14.8) [50-90] | 0.585 |
| **Smoking status – n (%)** |  |  |  |
| Smoker | 8 (5.67) | 24 (14.12) | **0.015** |
| Former smoker | 32 (22.70) | 28 (16.47) | 0.194 |
| Non smoker | 101 (71.63) | 118 (69.41) | 0.709 |
| **Body Mass Index – n (%)** |  |  |  |
| ≤29 | 100 (70.92) | 132 (77.65) | 0.192 |
| ≥30 | 41 (29.08) | 38 (22.35) | 0.192 |
| **SARS-CoV-2 serological status – n (%)** |  |  |  |
| Antibody-positive | 74 (52.48) | 134 (78.82) | **<0.001** |
| Antibody-negative | 64 (45.39) | 35 (20.59) | **<0.001** |
| Other | 3 (2.13) | 1 (0.59) |  |
| **Anti-SARS-CoV-2 vaccination status – n (%)** |  |  |  |
| 3 doses or 2 doses ≤ 120 days | 23 (16.31) | 66 (38.82) | **<0.001** |
| 1 or 2 doses ≥ 120 days or not vaccinated | 113 (80.14) | 99 (58.24) | **<0.001** |
| Other | 5 (3.55) | 5 (2.94) |  |
| **Comorbidities – n (%)** |  |  |  |
| Diabetes | 3 (2.13) | 6 (3.53) | 0.519 |
| Cardiovascular disease | 57 (40.43) | 61 (35.88) | 0.414 |
| Chronic kidney disease | 7 (4.96) | 9 (5.29) | 1.000 |
| Chronic liver disease | 4 (2.84) | 12 (7.06) | 0.061 |
| Chronic pulmonary disease | 19 (13.48) | 33 (19.41) | 0.173 |
| Immunocompromising conditions | 22 (15.60) | 39 (22.94) | 0.116 |
| **Symptoms at enrolment – n (%)** |  |  |  |
| Cough | 99 (70.21) | 118 (69.41) | 0.902 |
| Nasal congestion | 70 (49.65) | 69 (40.59) | 0.136 |
| Sore throat | 33 (23.40) | 69 (40.59) | **<0.001** |
| Feeling hot or feverish | 103 (73.05) | 99 (58.24) | **0.009** |
| Myalgia | 47 (33.33) | 54 (31.76) | 0.808 |
| Fatigue | 49 (34.75) | 75 (44.12) | 0.104 |
| Headache | 60 (42.55) | 60 (35.29) | 0.200 |
| Anosmia/ageusia | 41 (29.08) | 4 (2.35) | **<0.001** |
| Nausea/vomiting | 28 (19.86) | 11 (6.47) | **<0.001** |
| Diarrhoea | 15 (10.64) | 12 (7.06) | 0.314 |
| **Serum C-reactive protein level – n** | 140 | 161 |  |
| Mean (standard deviation) | 20.26 (28.66) | 14.29 (21.72) | **0.026** |

Comparing symptoms at enrolment by VOC, anosmia/ageusia (*p* <0.001), nausea/vomiting (*p* <0.001), and feeling feverish or hot (*p* <0.01) were significantly more frequent among patients infected with Delta, while sore throat (*p* <0.001) was significantly more frequent among patients infected with Omicron. Serological positivity to anti-SARS-CoV-2 antibodies (*p* <0.001) and complete primary COVID-19 vaccination series within 180 days of the enrolment or booster vaccination (*p* <0.001) were significantly more frequent among patients infected with Omicron. Table 2 shows the bivariate Cox regression of symptom resolution predictors by type of SARS-CoV-2 VOC. No predictors were associated with the time to symptom resolution in both SARS-CoV-2 VOC.

**Table 2.** Bivariate Cox regression of symptom resolution predictors by type of Variant of Concern.

| Predictor | Delta |  | Omicron |  |
| --- | --- | --- | --- | --- |
|  | Hazard ratio<br>(95% CI) | p value | Hazard ratio<br>(95% CI) | p value |
| Gender | 0.796<br>(0.569 - 1.113) | 0.182 | 0.836<br>(0.613 - 1.139) | 0.257 |
| Age | 1.0005<br>(0.984 - 1.017) | 0.952 | 0.996<br>(0.980 - 1.012) | 0.626 |
| Body Mass Index | 1.035<br>(0.716 - 1.495) | 0.855 | 1.171<br>(0.815 - 1.684) | 0.393 |
| SARS-CoV-2 serological status | 0.934<br>(0.667 - 1.307) | 0.690 | 0.823<br>(0.566 - 1.196) | 0.307 |
| Anti-SARS-CoV-2 vaccination status | 1.299<br>(0.827 - 2.040) | 0.257 | 0.906<br>(0.662 - 1.24) | 0.539 |
| Diabetes | 0.633<br>(0.34 - 1.179) | 0.150 | 1.19<br>(0.758 - 1.879) | 0.444 |
| Cardiovascular disease | 0.964<br>(0.686 - 1.354) | 0.831 | 0.852<br>(0.621 - 1.168) | 0.319 |
| Chronic kidney disease | 1.24<br>(0.577 - 2.665) | 0.581 | 1.125<br>(0.573 - 2.209) | 0.733 |
| Chronic liver disease | 2.417<br>(0.761 - 7.683) | 0.135 | 1.33<br>(0.738 - 2.401) | 0.341 |
| Chronic pulmonary disease | 0.778<br>(0.461 - 1.313) | 0.346 | 0.976<br>(0.666 - 1.431) | 0.902 |
| Immunocompromising conditions | 0.997<br>(0.597 - 1.664) | 0.989 | 0.802<br>(0.55 - 1.169) | 0.252 |

### Delta VOC

Baseline characteristics of 141 patients infected with Delta VOC by type of treatment are reported in Table 3. The main detected lineages were 34 AY.4 (24.1%), 33 AY.43 (23.4%), and 26 AY.122 (18.4%). 77 (54.6%) were male, median age was 65.7 years (IQR±15.4), 115 (78.8%) had at least one comorbidity, 74 (52.5%) were serum antibody-positive at the enrolment, and 23 (16.3%) received complete primary COVID-19 vaccination series within 180 days of the enrolment or booster vaccination.

**Table 3.** Baseline characteristics of the study population infected with Delta by type of treatment.

| Characteristic | Total<br>N = 141 | Sotrovimab<br>N = 44 | Bamlanivimab/<br>Etesevimab<br>N = 47 | Casirivimab/<br>Imdevimab<br>N = 50 |
| --- | --- | --- | --- | --- |
| **Sex (male) – n (%)** | 77 (54.61) | 23 (52.27) | 25 (53.19) | 29 (58.00) |
| **Age – median (interquartile range) [range]** | 65.7 (15.4) [50-92] | 65.8 (16.4) [50-90] | 68.6 (11.8) [50-92] | 63.2 (12) [50-89] |
| **Smoking status – n (%)** |  |  |  |  |
| Smoker | 8 (5.67) | 2 (4.45) | 4 (8.51) | 2 (4.00) |
| Former smoker | 32 (22.70) | 8 (18.18) | 11 (23.40) | 13 (26.00) |
| Non smoker | 101 (71.63) | 34 (77.27) | 32 (68.09) | 35 (70.00) |
| **Body Mass Index – n (%)** |  |  |  |  |
| ≤29 | 100 (70.92) | 29 (65.91) | 35 (74.47) | 36 (72.00) |
| ≥30 | 41 (29.08) | 15 (34.09) | 12 (25.53) | 14 (28.00) |
| **SARS-CoV-2 serological status – n (%)** |  |  |  |  |
| Antibody-positive | 74 (52.48) | 22 (50.00) | 30 (63.83) | 22 (44.00) |
| Antibody-negative | 64 (45.39) | 22 (50.00) | 16 (34.04) | 26 (52.00) |
| Other | 3 (2.13) | 0 | 1 (2.13) | 2 (4.00) |
| **Anti-SARS-CoV-2 vaccination status – n (%)** |  |  |  |  |
| 3 doses | 16 (11.35) | 6 (13.64) | 3 (6.38) | 7 (14.00) |
| 2 doses ≤ 120 days | 7 (4.96) | 2 (4.55) | 2 (4.26) | 3 (6.00) |
| 1 or 2 doses ≥ 120 days | 55 (39.01) | 16 (36.36) | 24 (51.06) | 15 (30.00) |
| Not vaccinated | 58 (41.13) | 18 (40.91) | 16 (34.04) | 24 (48.00) |
| Other | 5 (3.55) | 2 (4.55) | 2 (4.26) | 1 (2.00) |
| **Comorbidities – n (%)** |  |  |  |  |
| Diabetes | 3 (2.13) | 0 | 2 (4.26) | 1 (2.00) |
| Cardiovascular disease | 57 (40.43) | 18 (40.91) | 20 (42.55) | 19 (38.00) |
| Chronic kidney disease | 7 (4.96) | 1 (2.27) | 2 (4.26) | 4 (8.00) |
| Chronic liver disease | 4 (2.84) | 1 (2.27) | 1 (2.13) | 2 (4.00) |
| Chronic pulmonary disease | 19 (13.48) | 7 (15.91) | 5 (10.64) | 7 (14.00) |
| Immunocompromising conditions | 22 (15.60) | 6 (13.64) | 7 (14.89) | 9 (18.00) |
| **Symptoms at enrolment – n (%)** |  |  |  |  |
| Cough | 99 (70.21) | 30 (68.18) | 36 (76.60) | 33 (66.00) |
| Nasal congestion | 70 (49.65) | 20 (45.45) | 22 (46.81) | 28 (56.00) |
| Sore throat | 33 (23.40) | 11 (25.00) | 8 (17.02) | 14 (28.00) |
| Feeling hot or feverish | 103 (73.05) | 31 (70.45) | 36 (76.60) | 36 (72.00) |
| Myalgia | 47 (33.33) | 11 (25.00) | 16 (34.04) | 20 (40.00) |
| Fatigue | 49 (34.75) | 14 (31.82) | 15 (31.91) | 20 (40.00) |
| Headache | 60 (42.55) | 15 (34.09) | 15 (31.91) | 30 (60.00) |
| Anosmia/ageusia | 41 (29.08) | 13 (29.55) | 15 (31.91) | 13 (26.00) |
| Nausea/vomiting | 28 (19.86) | 6 (13.64) | 9 (19.15) | 13 (26.00) |
| Diarrhoea | 15 (10.64) | 1 (2.27) | 5 (10.64) | 9 (18.00) |
| **Serum C-reactive protein level – n** | 140 | 43 | 46 | 51 |
| Mean (standard deviation) | 20.26 (28.66) | 22.15 (33.04) | 25.27 (34.20) | 14.14 (15.78) |

No disease progression was recorded. An emergency department visit without hospitalisation was observed once in one patient in the casirivimab/imdevimab group. This visit was not deemed to be related to COVID-19.

The median time to symptom resolution was 7 days (95% CI 7 - 13) in the bamlanivimab/etesevimab group, 10 days (95% CI 7 - 14) in the sotrovimab group, and 10 days (95% CI 7 - 15) in the casirivimab/imdevimab group, not differing significantly among the overall groups of treatment (Log-rank Chi-square 0.22, *p* 0.895) and for each comparison between treatment groups, namely bamlanivimab/etesevimab with casirivimab/imdevimab (Log-rank Chi-square 0.08, *p* 0.776), sotrovimab with casirivimab/imdevimab (Log-rank Chi-square 0.40, *p* 0.527), and bamlanivimab/etesevimab with sotrovimab (Log-rank Chi-square 0.01, *p* 0.92). Figure 1 shows the survival time to symptom resolution by type of treatment in the Delta study population. The Cox regression analysis confirmed the non-significantly different effects upon the time to symptom resolution between casirivimab/imdevimab (reference standard according to the original trial protocol) and both bamlanivimab/etesevimab and sotrovimab (HR 1.052 and HR 1.097, 95% CI 0.704 - 1.573 and 0.729 - 1.652, *p* 0.805 and 0.657, respectively).

**Figure 1.**
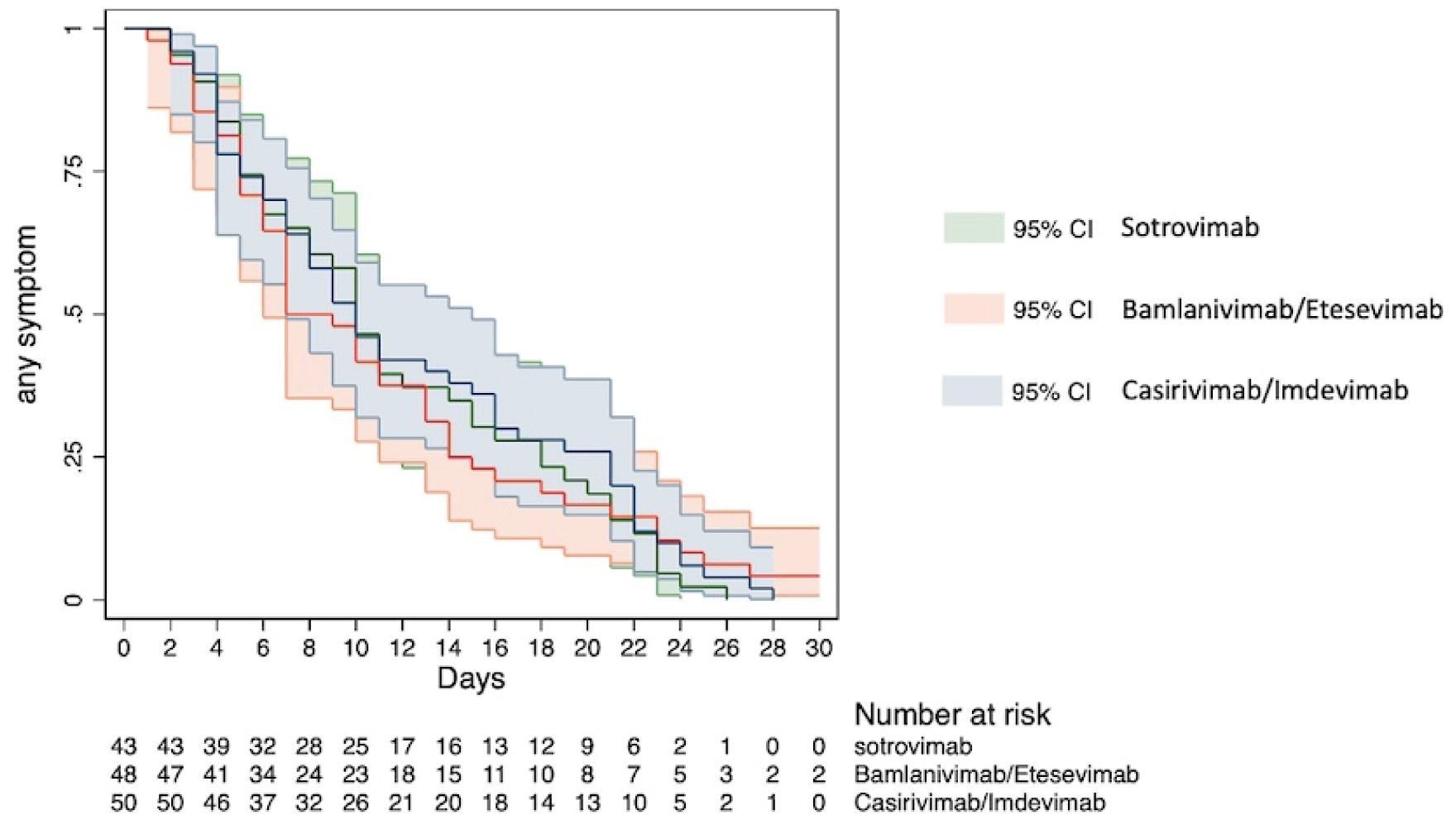
Survival time to symptom resolution by type of treatment in the study population infected with Delta.

### Omicron VOC

Baseline characteristics of 170 patients infected with Omicron VOC by type of treatment are reported in Table 4. The detected lineages were 137 (80.6%) BA.1 and 33 (19.4%) BA.1.1. 101 (59.4%) were male, median age was 64.5 years (IQR±14.8), 135 (79.4%) had at least one comorbidity, 134 (78.8%) were serum antibody-positive at the enrolment, and 66 (38.8%) received complete primary COVID-19 vaccination series within 180 days of the enrolment or booster vaccination.

**Table 4.** Baseline characteristics of the study population infected with Omicron by type of treatment.

| Characteristic | Total<br>N = 170 | Sotrovimab<br>N = 61 | Bamlanivimab/<br>Etesevimab<br>N = 57 | Casirivimab/<br>Imdevimab<br>N = 52 |
| --- | --- | --- | --- | --- |
| **Sex (male) – n (%)** | 101 (59.41) | 36 (59.02) | 30 (52.63) | 35 (67.31) |
| **Age – median (interquartile range) [range]** | 64.5 (14.8) [50-90] | 64.2 (15) [50-90] | 64.8 (14.6) [50-86] | 65.3 (14.8) [50-86] |
| **Smoking status – n (%)** |  |  |  |  |
| Smoker | 24 (14.12) | 6 (9.84) | 11 (19.30) | 7 (13.46) |
| Former smoker | 28 (16.47) | 9 (14.75) | 11 (19.30) | 8 (15.38) |
| Non smoker | 118 (69.41) | 46 (75.41) | 35 (61.40) | 37 (71.15) |
| **Body Mass Index – n (%)** |  |  |  |  |
| ≤29 | 132 (77.65) | 53 (86.89) | 42 (73.68) | 37 (71.15) |
| ≥30 | 38 (22.35) | 8 (13.11) | 15 (26.32) | 15 (28.85) |
| **SARS-CoV-2 serological status – n (%)** |  |  |  |  |
| Antibody-positive | 134 (78.82) | 45 (73.77) | 45 (78.95) | 44 (84.62) |
| Antibody-negative | 35 (20.59) | 16 (26.23) | 11 (19.30) | 8 (15.38) |
| Other | 1 (0.59) | 0 | 1 (1.75) | 0 |
| **Anti-SARS-CoV-2 vaccination status – n (%)** |  |  |  |  |
| 3 doses | 62 (36.47) | 24 (39.34) | 19 (33.33) | 19 (36.54) |
| 2 doses ≤ 120 days | 4 (2.35) | 2 (3.28) | 1 (1.75) | 1 (1.92) |
| 1 or 2 doses ≥ 120 days | 57 (33.53) | 16 (26.23) | 22 (38.60) | 19 (36.54) |
| Not vaccinated | 42 (24.71) | 18 (29.51) | 13 (22.81) | 11 (21.15) |
| Other | 5 (2.94) | 1 (1.64) | 2 (3.51) | 2 (3.85) |
| **Comorbidities – n (%)** |  |  |  |  |
| Diabetes | 6 (3.53) | 2 (3.28) | 2 (3.51) | 2 (3.85) |
| Cardiovascular disease | 61 (35.88) | 18 (29.51) | 17 (29.82) | 26 (50.00) |
| Chronic kidney disease | 9 (5.29) | 4 (6.56) | 2 (3.51) | 3 (5.77) |
| Chronic liver disease | 12 (7.06) | 4 (6.56) | 5 (8.77) | 3 (5.77) |
| Chronic pulmonary disease | 33 (19.41) | 11 (18.03) | 15 (26.32) | 7 (13.46) |
| Immunocompromising conditions | 39 (22.94) | 17 (27.87) | 11 (19.30) | 11 (21.15) |
| **Symptoms at enrolment – n (%)** |  |  |  |  |
| Cough | 118 (69.41) | 42 (68.85) | 37 (64.91) | 39 (75.00) |
| Nasal congestion | 69 (40.59) | 28 (45.90) | 25 (43.86) | 16 (30.77) |
| Sore throat | 69 (40.59) | 22 (36.07) | 27 (47.37) | 20 (38.46) |
| Feeling hot or feverish | 99 (58.28) | 37 (60.66) | 32 (56.14) | 30 (57.69) |
| Myalgia | 54 (31.76) | 20 (32.79) | 18 (31.58) | 16 (30.77) |
| Fatigue | 75 (44.12) | 31 (50.82) | 20 (35.09) | 24 (46.15) |
| Headache | 60 (35.29) | 23 (37.70) | 20 (35.09) | 17 (32.69) |
| Anosmia/ageusia | 4 (2.35) | 1 (1.64) | 2 (3.51) | 1 (1.92) |
| Nausea/vomiting | 11 (6.47) | 4 (6.56) | 5 (8.77) | 2 (3.85) |
| Diarrhoea | 12 (7.06) | 5 (8.20) | 4 (7.02) | 3 (5.77) |
| **Serum C-reactive protein level – n** | 161 | 57 | 56 | 48 |
| Mean (standard deviation) | 14.29 (21.72) | 12.65 (15.97) | 17.19 (31.07) | 12.87 (12.55) |

Two of 53 in the bamlanivimab/etesevimab group (3.8%) had disease progression leading to hospitalisation and no disease progression was recorded in the casirivimab/imdevimab and sotrovimab groups. The primary reasons for the two hospitalisations were deemed to be related to COVID-19. Both patients admitted to hospital were serum antibody-negative at enrolment and underwent non-invasive mechanical ventilation at hospital admission. One of these patients, a man aged 71-75 who received three doses of SARS-CoV-2 vaccine and was affected by non-Hodgkin lymphoma under active chemotherapy and chronic heart failure, died 12 days after the symptom onset, 10 days after the administration of bamlanivimab/etesevimab, and 4 days after the hospitalisation. The other patient, a man aged 66-70 who was not vaccinated against SARS-CoV-2 and was affected by obesity (body mass index, 31) and type 2 diabetes, was admitted 7 days after the symptom onset and 4 days after the administration of bamlanivimab/etesevimab; the length of his hospital stay was 22 days, including non-invasive mechanical ventilation for 13 days and low-flow oxygen therapy for 8 days. All-cause mortality through day 28 was the same as the one through day 14.

An emergency department visit without hospitalisation was observed once in one patient in the bamlanivimab/etesevimab group. This visit was not deemed to be related to COVID-19.

The median time to symptom resolution was 12 days (95% CI 8 - 14) in the bamlanivimab/etesevimab group, 12 days in the casirivimab/imdevimab group (95% CI 9 - 16), and 7 days in the sotrovimab group (95% CI 6 - 9), turning out to be 5 days shorter in the sotrovimab group compared to both bamlanivimab/etesevimab and casirivimab/imdevimab groups (HR 0.526 and HR 0.451, 95% CI 0.359 - 0.77 and 95% CI 0.303 - 0.669, *p* 0.001 and 0.0001, respectively). Figure 2 shows the survival time to symptom resolution by type of treatment in the Omicron study population. In each of the assessed subgroups (SARS-CoV-2 serological and vaccination status), sotrovimab showed a significantly shorter time to symptom resolution compared to bamlanivimab/etesevimab and casirivimab/imdevimab, as reported in Table 5.

**Table 5.** Cox regression to assess the difference between treatment effects upon the time to symptom resolution in selected subgroups of interest in the study population infected with Omicron.

| Subgroup | Sotrovimab | Bamlanivimab/etesevimab |  | Casirivimab/imdevimab |  |
| --- | --- | --- | --- | --- | --- |
|  | Hazard ratio | Hazard ratio (95% CI) | p value | Hazard ratio (95% CI) | p value |
| **SARS-CoV-2 serological status** |  |  |  |  |  |
| Antibody-negative | 1 | 0.345 (0.158 - 0.753) | **0.008** | 0.415 (0.177 -0.973) | **0.043** |
| Antibody-positive | 1 | 0.398 (0.221 - 0.714) | **0.002** | 0.318 (0.175 - 0.575) | **<0.001** |
| **Anti-SARS-CoV-2 vaccination status** |  |  |  |  |  |
| 1 or 2 doses ≥ 120 days or not vaccinated | 1 | 0.472 (0.289 - 0.772) | **0.003** | 0.495 (0.298 - 0.821) | **0.006** |
| 3 doses or 2 doses ≤ 120 days | 1 | 0.504 (0.284 - 0.894) | **0.019** | 0.346 (0.194 - 0.62) | **<0.001** |

**Figure 2.**
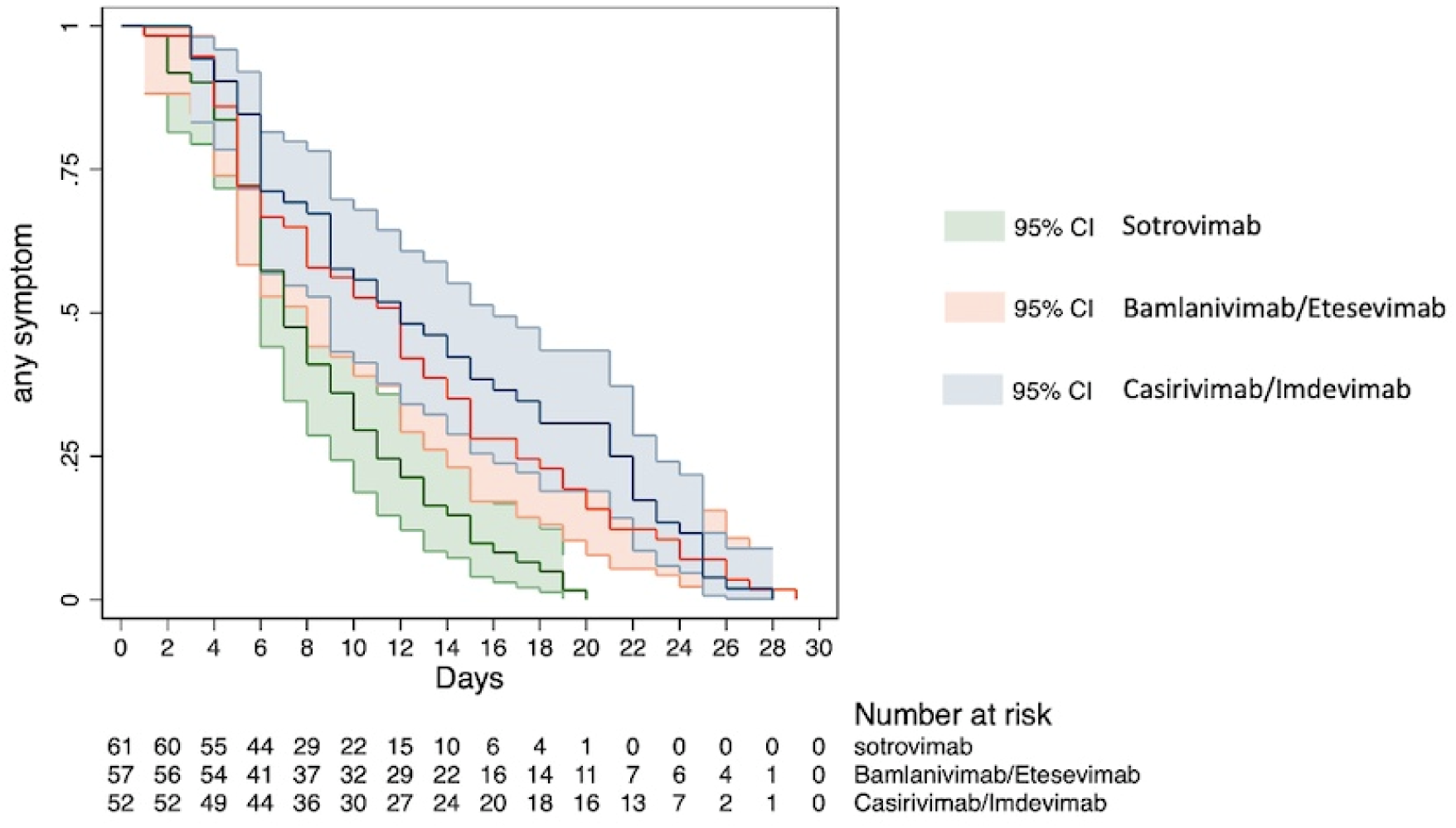
Survival time to symptom resolution by type of treatment in the study population infected with Omicron.

## Discussion

During the SARS-CoV-2 pandemic, the paradigm of discovering and implementing mAb and antiviral treatments based on the randomised controlled trials has lagged significantly behind the new evidence coming from in-vitro studies, which has driven clinical recommendations causing ethical dilemmas on the continuation of ongoing trials. At the time of approving the MANTICO trial protocol (November 2021), casirivimab/imdevimab, bamlanivimab/etesevimab, and, later, sotrovimab were the only therapies recommended by the COVID-19 treatment guidelines for outpatients with mild to moderate COVID-19 at high risk of progressing to severe disease. Delta was the SARS-CoV-2 dominant VOC worldwide and the selection of the study mAbs was based on their in-vitro activity against the circulating variants and on the existing evidence of their clinical efficacy. After mid-December 2021, the Omicron VOC has been spreading worldwide, rapidly becoming the dominant VOC. Preliminary in-vitro studies on Omicron demonstrated numerous mutations in the gene encoding the spike protein, predicting a markedly reduced susceptibility to bamlanivimab/etesevimab and casirivimab/imdevimab [3-5]. According to these findings, FDA and AIFA have revised the emergency use authorization for bamlanivimab/etesevimab and casirivimab/imdevimab, halting their use, in line with the National Institutes of Health (NIH) COVID-19 Treatment Guidelines Panel, which advised against the use of these mAbs due to reduced activity against Omicron and because real-time testing to identify rare, non-Omicron variants is not readily available [15]. Therefore, the study enrolment in a real-life outpatient setting was prematurely discontinued for possible futility, after the inclusion of barely one fourth of the predefined sample size. Nevertheless, the recruitment timeframe provided a unique opportunity to collect data on the clinical efficacy of bamlanivimab/etesevimab, casirivimab/imdevimab, and sotrovimab in patients infected with Omicron.

Overall, the three treatment groups appeared to be balanced with respect to the predictors of outcomes in both Delta and Omicron population, as expected under the randomised allocation design. As reported by previous studies, patients infected with Omicron were more likely than those infected with Delta to present with symptoms limited to the upper respiratory tract and to have pre-existing immunity, considering that Omicron is better equipped than Delta to infect people with pre-existing immunity [16].

Considering the time to symptom resolution, no differences in the effect between treatment groups was found in Delta infections, whereas sotrovimab showed a significant benefit compared to bamlanivimab/etesevimab and casirivimab/imdevimab in patients infected with Omicron BA.1 and BA.1.1. This benefit was consistent across all Omicron subgroups, regardless of the SARS-CoV-2 serology and vaccination status, confirming the preliminary in-vitro evidence on the mAbs activity against Omicron BA.1 and BA.1.1 [3-5].

The disease progression was recorded in two patients infected with Omicron, who were both randomised to receive bamlanivimab/etesevimab. The absence of COVID-19 progression in two treatment groups (casirivimab/imdevimab and sotrovimab) in the Omicron study population, as well as in all three treatment groups in the Delta study population, could be influenced by the small sample size, the lower intrinsic-severity of Omicron, the high vaccination rate in Italy, and the prioritization of the booster vaccination for the elderly [17]. Nevertheless, these findings are consistent with recent in-vitro data showing that all study treatments were active against Delta and both casirivimab/imdevimab and sotrovimab retained a residual neutralizing activity against Omicron, whereas bamlanivimab/etesevimab did not neutralize Omicron [18-20]. Additional clinical studies with an adequate sample size are required to determine whether casirivimab/imdevimab and sotrovimab are indeed effective in preventing COVID-19 progression due to Omicron infection. Should the role of casirivimab/imdevimab in preventing severe COVID-19 due to Omicron infections be confirmed, this mAb could represent a readily-available and well-tolerated treatment option in case of shortages of mAbs supplies and contraindication to other early COVID-19 treatments.

The MANTICO trial provides the first data on the clinical efficacy of bamlanivimab/etesevimab, casirivimab/imdevimab, and sotrovimab against Omicron VOC. There is an urgent need for adaptive clinical trials comparing anti-SARS-CoV-2 treatments by VOC to promptly inform recommendations for the management of early COVID-19.

## Data Availability

Data are available upon reasonable request to the corresponding author. Individual records will be anonymised and shared in STATA format.

